# Knowledge and practice toward pelvic floor health: a cross-sectional study protocol within the medical staff of national Athletics teams

**DOI:** 10.1101/2025.09.16.25335684

**Authors:** Giagio Silvia, Adami Paolo Emilio, Bermon Stephane, Rial-Rebullido Tamara, Turolla Andrea, Pillastrini Paolo, Garrandes Frederic

## Abstract

**Objectives:** This study will aim to assess the knowledge and practices related to pelvic floor (PF) health within the medical staff of national Athletics teams.

Secondary objectives will be to a) explore differences in PF health knowledge across individual, professional, and contextual characteristics; b) examine whether individual clinical practices and team-level approaches toward PF health will vary across subgroups.

**Methods:** This study will be an observational, cross-sectional design using a web-based survey, conducted during the World Athletics season from September 2025 to September 2026. Participants will include healthcare professionals who are part of the medical staff of national Athletics teams, such as physicians, physiotherapists, nurses, nutritionists, and other allied health professionals. The survey will assess demographic and professional characteristics, basic knowledge and practices related to PF health. The questionnaire will undergo expert review and pilot testing before implementation. Participation will be voluntary and anonymous, with responses securely stored. Data will be analyzed using descriptive statistics and comparative tests to explore group differences.

**Conclusions:** Findings will provide insight into how medical staff address PF health within elite Athletics, highlighting potential gaps in knowledge and clinical practice. Data will inform educational initiatives and policy recommendations, supporting the integration of PF health into elite athlete care and sports medicine training programs.

**Revision summary:** The data collection period has been extended from the Tokyo 2025 World Athletics Championships (13–21 September 2025) to the World Athletics Road Running Championships in Copenhagen (19–20 September 2026), resulting in an overall data collection period of one year.

No other changes have been made to the study objectives, eligibility criteria, survey content, recruitment procedures, outcome measures, or statistical analysis plan..

## BACKGROUND

Over the past decade, pelvic floor (PF) health has gained increasing attention in sports medicine. Most research and international initiatives primarily focused on athletes, exploring epidemiology, awareness and potential interventions [1–6]. In particular, the literature highlighted a high prevalence of PF dysfunction in high-impact and high-intensity sports, such as Athletics, with consistent evidence linking these symptoms to reduced athletic performance and quality of life [7–9].

While these findings have advanced knowledge on PF in athletes, research on health professionals’ perspectives on PF health in sports medicine remains limited. Yet, elite sports rely on multidisciplinary medical staff, where health professionals are fundamental in monitoring athlete health, identifying potential concerns, and managing medical conditions. The few available studies, conducted outside the elite context, found poor awareness, inconsistent screening practices, and an overall lack of standardized approaches [10]. Furthermore, the Periodic Health Evaluation (PHE), a well-known component of athlete health monitoring, does not currently include PF domain [11,12], leaving this area largely overlooked [13].

This observational, cross-sectional, web-based survey will aim to address this gap by assessing the knowledge and practices within the medical staff of national Athletics teams toward PF health. This study is part of a broader World Athletics’ initiative. Findings will provide evidence to guide educational interventions, improve screening strategies, and inform policy recommendations for integrating PF health into elite athlete care and sports medicine training programs.

### OBJECTIVES

The primary objectives of this study will be to assess the knowledge and practices of national Athletics’ teams medical staff toward PF health.

Secondary objectives will be a) to assess whether PF health knowledge differs across individual, professional and contextual characteristics; b) to assess whether the integration of PF health into clinical practice at the individual level varies according to individual, professional, and contextual characteristics; c) to investigate whether team-level practices toward PF health differ across contextual characteristics.

## METHODS

### Study design

This study will be an observational, cross-sectional design using a web-based survey, over a one-year period. For the reporting, the Checklist for Reporting Results of Internet E-Surveys (CHERRIES) and the Strengthening the Reporting of Observational Studies in Epidemiology (STROBE) will be used. Ethical approval has been obtained from the Bioethics Committee of Bologna University (Prot. N. 0184744). All participants will provide informed consent before participation, and responses will be anonymized to ensure confidentiality.

### Participants

#### Inclusion criteria

The study population will include the medical staff of national Athletics teams traveling with athletes to World Athletics Series events between September 2025 and September 2026.

Eligible events were identified according to the official World Athletics competition calendar and include the World Athletics Ultimate Championship. The complete calendar is available at the following website link: https://worldathletics.org/competition/calendar-results?competitionGroupId=3806).

In particular, the following eight events will be considered: World Athletics Championships - Tokyo (JPN) 13-21 September 2025; 46th World Athletics Cross Country Championships - Tallahassee, FL (USA), 10 January 2026); World Athletics Indoor Championships - Toruń (POL), 20-22 March 2026; Caixa World Athletics Race Walking Team Championships Brasilia 26 - Brasilia (BRA), 12 April 2026; World Athletics Relays - Gaborone (BOT), 2-3 May 2026; World Athletics U20 Championships - Eugene, OR (USA), 05-09 August 2026; World Athletics Ultimate Championship - Budapest (HUN), 11-13 September 2026; World Athletics Road Running Championships – København, 19-20 September 2026

Professionals will be defined according to the World Health Organization (WHO) classification of health professionals (**Supplementary File 1**).

#### Exclusion criteria

Individuals with professional title outside the WHO-defined health professional classifications will be excluded from the study.

### Research team

The research team comprised two women and five men, including clinicians and researchers from various specialties and nationalities representing Italy, Spain, France and the USA. The team’s expertise spanned research methodology, sports medicine, musculoskeletal/sport physiotherapy, PF physiotherapy, and exercise physiology.

### Survey development

Survey items will be developed by the research team based on previous literature [10,14–20] with adaptations. Considerations will be made to ensure clarity, medical relevance, and sensitivity to cultural differences. The survey will be also designed considering time-efficiency to maximize response rates.

### Questionnaire validation and pilot testing

After drafting the initial version of the survey, a panel of experts in research methodology will review it to assess the structure, organization, logical flow, appropriateness of question formats, and overall accuracy [21]. Feedback from this phase will be used to refine the survey’s methodological aspects before moving to content validation. Then, a pilot test will be conducted with health professionals, including physiotherapists, sports medicine practitioners, and pelvic floor specialists. This phase will evaluate the survey’s clarity, content relevance, acceptability, and response format [21]. Any necessary modifications will be made to improve the comprehensibility and applicability of the questionnaire before its finalization. Data from the pilot phases will not be included in the final analysis. Given that this survey is designed specifically for this study, it will not undergo formal psychometric validation.

## Survey content

The survey will mainly consist of three sections:

1. ***Participants’ characteristics*** This section will collect main demographic and professional information, including age, sex, national staff affiliation of Athletics medical teams, professional role, and years of experience.
2. ***Knowledge*** This section will assess participants’ baseline knowledge of PF health. It will include a cluster of multiple-choice questions, each with a single correct answer. Correct responses will be assigned 1 point, while incorrect responses will receive 0 points. A cumulative knowledge score will be calculated, with higher scores indicating greater knowledge. Examples of questions will cover: Basic anatomy and physiology of the PF; PF function in sport; Terminology and definitions of common PF dysfunctions; Awareness of subgroups at higher risk.
3. ***Practices***. This section will investigate both individual-level and team-level practices. It will include Yes/No questions and multiple-choice items. No cumulative score will be assigned. Individual-level practices will assess whether professionals discuss PF health with athletes, feel comfortable addressing the topic, provide education, and perform basic screening. In addition, perceived preparedness will be evaluated on a 0–10 Likert scale. Team-level practices will explore how PF health is addressed within national Athletics team medical staff.

An open-ended question will allow respondents to provide additional comments, perspectives, or suggestions regarding PF health in elite Athletics. See **Supplementary File 2** for detailed information.

The survey will be available in English and will be hosted on SurveyMonkey (SurveyMonkey, Palo Alto, California).

### Procedure

During each event, the medical staff of national Athletics teams will be invited to participate by scanning a QR code distributed at the official medical conference and medical meeting. In addition, members of the World Athletics Health and Science Department (PEA, FG, SB) will further engage with professionals on site, providing information, clarifying questions, and supporting participation throughout the competition days. Data will be collected exclusively during the championship period. Participation will be voluntary, with no incentives provided.

Participants will have the option to withdraw at any time before survey submission. Responses can be reviewed and modified before final submission. All data will be securely stored on an encrypted computer, with access restricted to the research team. Participants will be ensured that their identities will remain confidential and will not be disclosed to the investigators [21]. Upon survey completion, professionals will be able to download an educational leaflet on PF health developed by the World Athletics Health and Science Department (SG, FG, PEA, SB).

### Sample size calculation

The required sample size will be estimated using the formula for proportions with 95% confidence level, p=0.5 and 5% margin of error. A finite population correction will be applied, considering the number of eligible professionals recorded in the official World Athletics registry available at the time of competition.

### Statistical methods

Survey responses will be exported from SurveyMonkey to Excel and reviewed for completeness. Only fully completed responses will be included in the analysis.

Descriptive statistics will be used to summarize participant characteristics and survey responses. Categorical variables will be reported as frequencies and percentages. Continuous variables will be presented as means and standard deviations or medians and interquartile ranges, depending on distribution, which will be assessed using the Shapiro–Wilk test.

Differences in knowledge scores across subgroups (e.g., sex, professional title, WHO region) will be assessed using parametric (e.g., t-test, ANOVA) or non-parametric tests (e.g., Mann–Whitney U, Kruskal–Wallis), and depending on data distribution. Multivariable linear or logistic regression models will be used to identify factors independently associated with knowledge scores, screening practice, adjusting for relevant covariates. A statistical significance level of p<0.05 will be used. Data analyses will be performed using STATA 19 SE, and results will be reported with corresponding confidence intervals and effect sizes where applicable.

Open-ended comments will be reviewed for common themes.

## Data Availability

Not applicable at this stage.

## Acknowledgements

Not applicable.

## SUPPLEMENTARY FILE 1

| Occupation group | ISCO code | Definition | Examples of occupations classified here | Notes |
| --- | --- | --- | --- | --- |
| **HEALTH PROFESSIONALS** |  |  |  |  |
| Health professionals study, advise on or provide preventive, curative, rehabilitative and promotional health services based on an extensive body of theoretical and factual knowledge in diagnosis and treatment of disease and other health problems. They may conduct research on human disorders and illnesses and ways of treating them, and supervise other workers. The knowledge and skills required are usually obtained as the result of study at a higher educational institution in a health-related field for a period of 3–6 years leading to the award of a first degree or higher qualification. |  |  |  |  |
| Generalist medical practitioners | 2211 | Generalist medical doctors (including family and primary care doctors) diagnose, treat and prevent illness, disease, injury, and other physical and mental impairments and maintain general health in humans through application of the principles and procedures of modern medicine. They plan, supervise and evaluate the implementation of care and treatment plans by other health care providers. They do not limit their practice to certain disease categories or methods of treatment, and may assume responsibility for the provision of continuing and comprehensive medical care to individuals, families and communities. | Medical doctor (general), Medical officer (general), Physician (general), General practitioner, Family medical practitioner, Primary health care physician, District medical doctor, Resident medical officer specializing in general practice, Township medical officer, Station medical officer, Specialist in family medicine, Medical officer | Occupations included in this category require completion of a university-level degree in basic medical education plus postgraduate clinical training or equivalent. Medical interns who have completed their university education in basic medical education and are undertaking postgraduate clinical training are included here. Although in some countries 'general practice' and 'family medicine' may be considered as medical specializations, these occupations should always be classified here. |

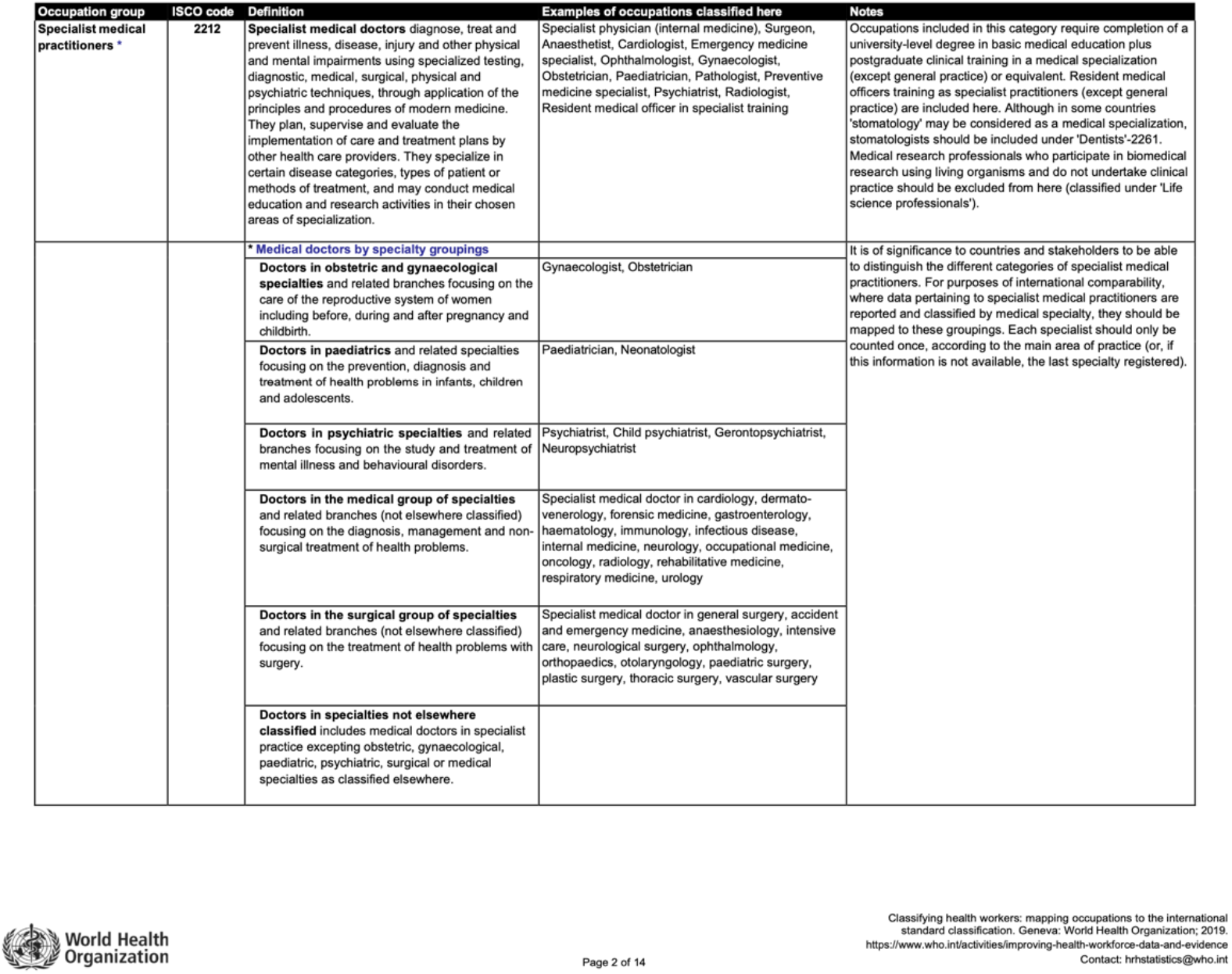

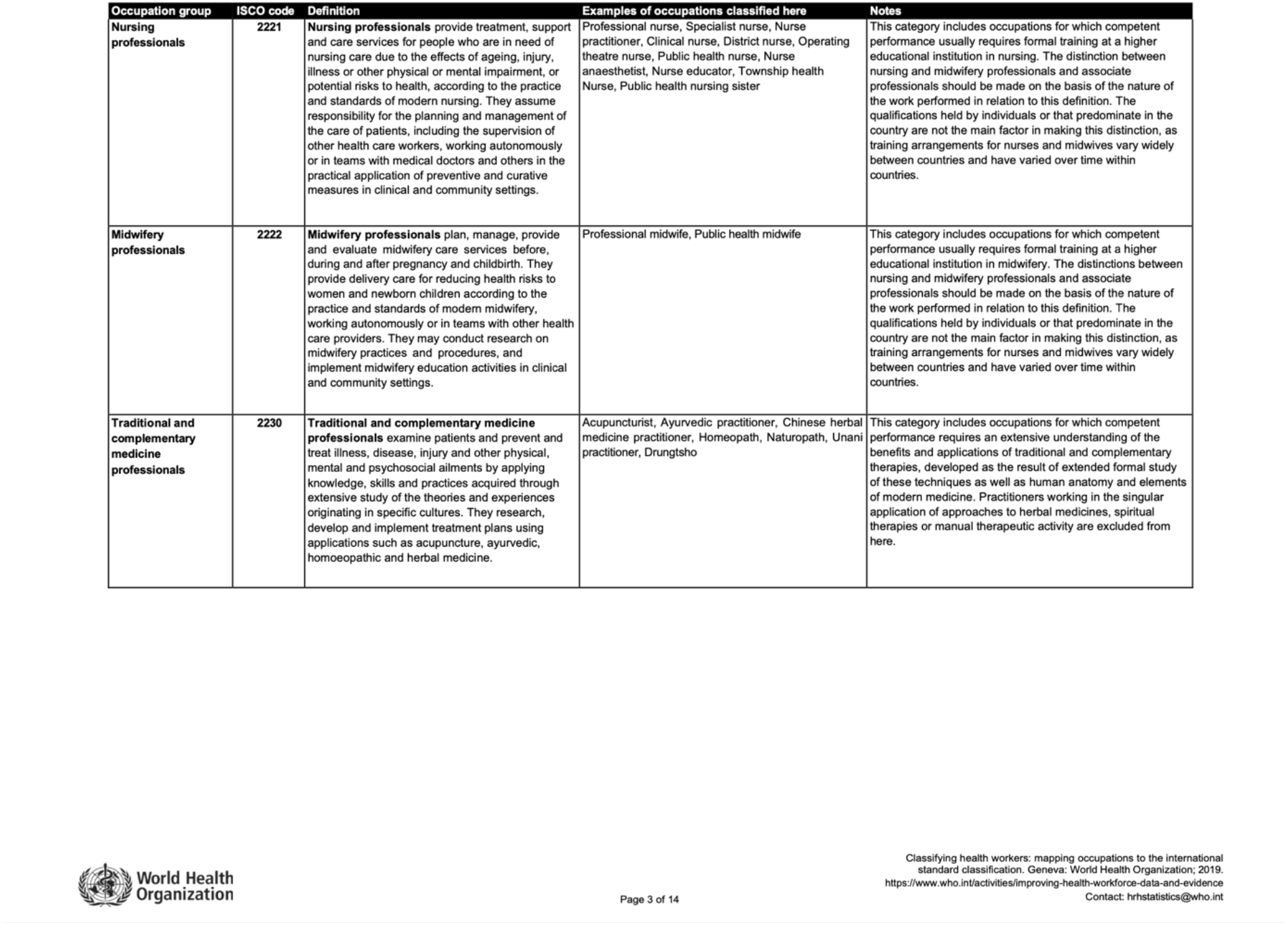

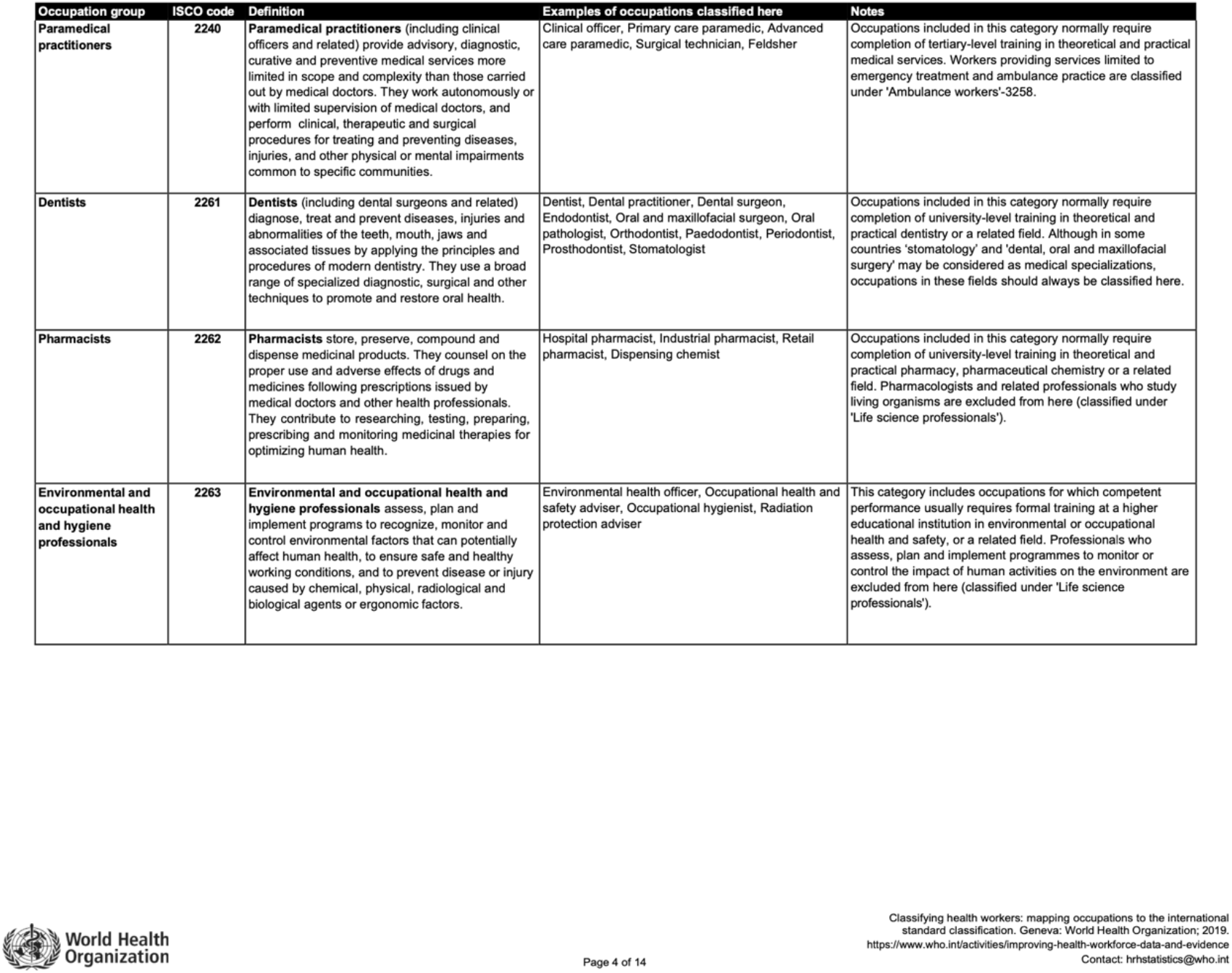

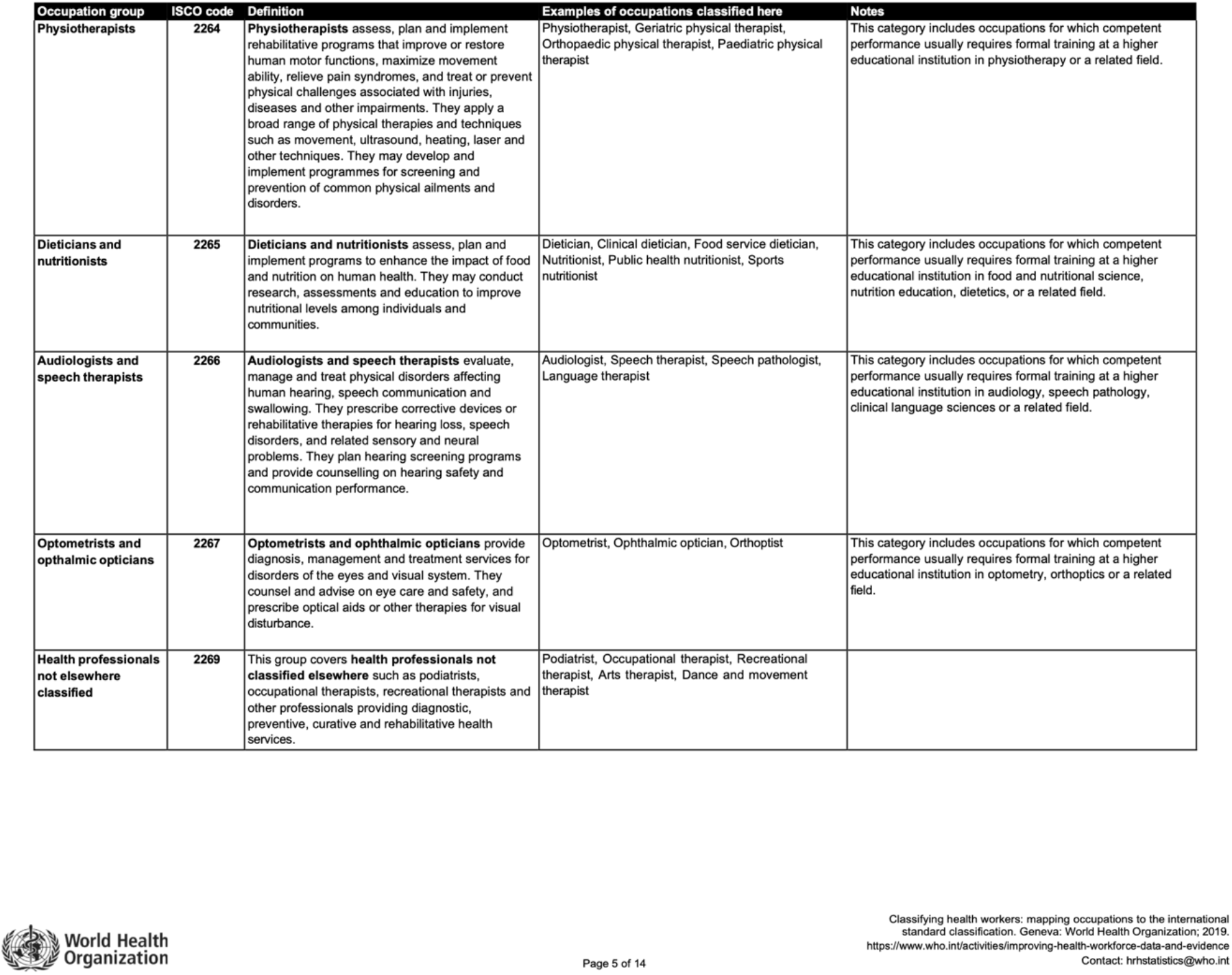

## SUPPLEMENTARY FILE 2

### DEMOGRAPHIC INFORMATION - SECTION 1

1. What is your **sex**?
  □ Male
  □ Female
  □ I prefer not to say
2. How old are you? (years, numbers only)
3. Which **national Athletics’ team medical staff** are you currently working with?
  □ *List*
4. What is your **professional role (job title)**?
  □ Sports medicine physician
  □ Other physician (all other specialties, e.g. general practitioner, cardiologist)
  □ Physiotherapist
  □ Other (not included in the options above)
5. How many years of experience do you have working in **elite Athletics**?
  □ 5 years or less
  □ 6–10 years
  □ More than 10 years

### KNOWLEDGE - SECTION 2

This section aims to assess basic knowledge of pelvic floor health. Please indicate the correct answer.

1. **In your opinion, which number shows the pelvic floor location correctly on the body figure?**
  a. 1
  b. 2
  c. 3
  d. 4
  e. 5
2. **Which of the following is a primary function of the pelvic floor?**
  a. Adapting abdominal wall during exertion
  b. Supporting pelvic organs and maintaining continence
  c. Enhancing spinal flexibility, providing core stabilization
  d. Increasing intra-abdominal pressure for maximal strength output
  e. All of the above
3. **Which of the following combinations are symptoms of pelvic floor dysfunction?**
  a. Urgency urinary incontinence and pelvic cramping
  b. Urinary retention and tight pelvic floor muscles
  c. Pelvic heaviness and pelvic cramping
  d. Urinary and anal incontinence
  e. All of the above
4. **Which of these athlete groups can have pelvic floor problems?**
  a. Only female athletes with a history of pregnancy
  b. Mainly elite female athletes involved in endurance sports
  c. Athletes performing only high-impact sports at professional level
  d. Male and female athletes with a history of pelvic trauma or surgery
  e. Both male and female athletes, including adolescents, regardless of sports
5. **According to the literature, what is the average prevalence of urinary incontinence among elite female athletes?**
  a. Less than 5%
  b. 5 – 30%
  c. 30 – 60%
  d. More than 60%
  e. I do not know/ I am not sure
6. **Which of the following is the first-line treatment for pelvic floor dysfunction in athletes?**
  a. Use of compression garments during training
  b. Individualized pelvic floor muscle training
  c. Medications including systemic anti-inflammatory drugs (NSAIDs)
  d. Rest, and avoidance of activities that trigger symptoms for at least 2 weeks
  e. All the above

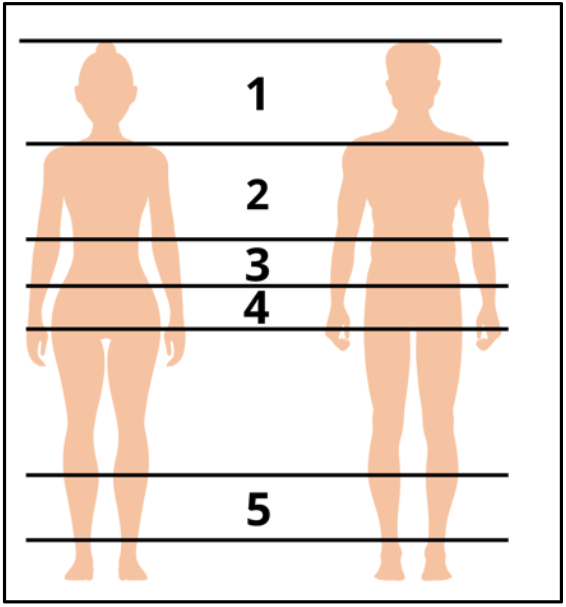

### PRACTICES – SECTION 3

#### GENERAL BACKGROUND

Please answer the following questions with: Yes/No

|  | **No** | **Yes** |
| --- | --- | --- |
| Have you received any formal training or education in pelvic floor health? |  |  |
| Have you previously or ever searched for information about pelvic floor health? |  |  |
| Are you personally interested in pelvic floor health? |  |  |

#### INDIVIDUAL PRACTICES IN ELITE ATHLETICS

These questions refer to practices that you personally adopt in your role within elite Athletics.

**Answer: Yes / No**

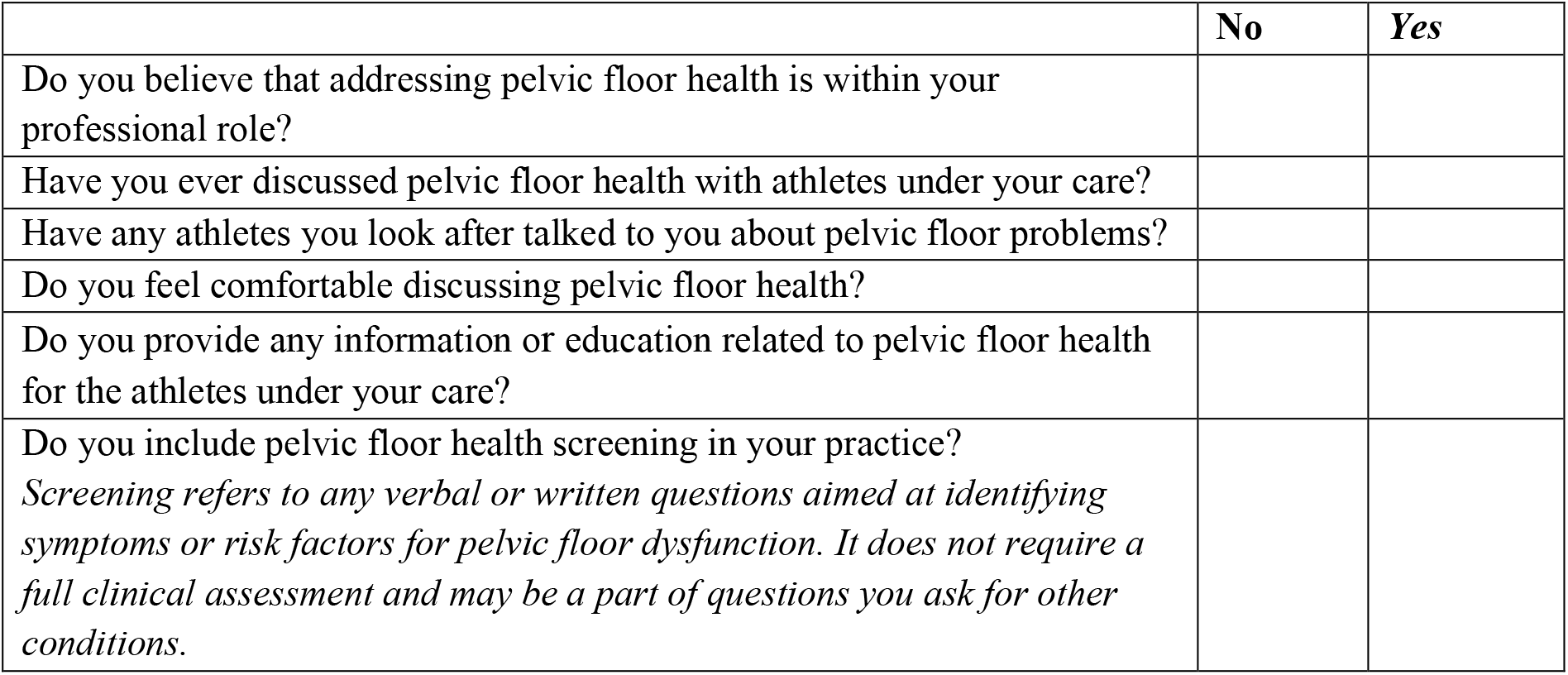

|  | No | Yes |
| --- | --- | --- |
| Do you believe that addressing pelvic floor health is within your professional role? |  |  |
| Have you ever discussed pelvic floor health with athletes under your care? |  |  |
| Have any athletes you look after talked to you about pelvic floor problems? |  |  |
| Do you feel comfortable discussing pelvic floor health? |  |  |
| Do you provide any information or education related to pelvic floor health for the athletes under your care? |  |  |
| Do you include pelvic floor health screening in your practice?<br>*Screening refers to any verbal or written questions aimed at identifying symptoms or risk factors for pelvic floor dysfunction. It does not require a full clinical assessment and may be a part of questions you ask for other conditions.* |  |  |

Which categories of athletes do you screen for pelvic floor health? (*Select all that apply, multiple choice)*

1. None, I do not screen any athletes
2. All athletes
3. All female athletes
4. Females with a history of pregnancy
5. Athletes with trauma or injury in the pelvic area, regardless of sex
6. Other (please specify)

Which methods do you use for screening for pelvic floor health? (*Select all that apply, multiple choice)*

1. None, I do not screen for pelvic floor health
2. Standardized tools/questionnaires specific to pelvic floor health
3. Informal discussion without standardized tools/questionnaires specific to pelvic floor health
4. Other (please specify)

On a scale from 0 to 10, how prepared do you feel to address pelvic floor health within your current professional role?

0 = Not at all prepared, 10 = Fully prepared

### MEDICAL STAFF/TEAM PRACTICES

These questions refer to practices that are implemented at the team or organizational level toward pelvic floor health.

How is pelvic floor health addressed within your national Athletics team medical staff? (*Select all that apply, multiple choice)*

1. I do not know
2. It is openly discussed
3. There is no structured way to discuss or report it
4. It is discussed only upon athlete request
5. It is referred to external pelvic floor specialists
6. Other (please specify):_____________

Does your national medical team have an identified internal or external consultant specializing in pelvic floor health (e.g., a pelvic floor physiotherapist, urologist, or uro/gynecologist)?

1. No
2. Yes
3. I do not know/I am not sure

### OPTIONAL OPEN-ENDED QUESTION

If you would like to add any comments, please do so here:__________________

## Notes

### Competing Interest Statement

The authors have declared no competing interest.

### Author Declarations

Bioethics Committee of the University of Bologna, Italy (Prot. N. 0184744)

### Summary of Updates

The data collection period has been extended from the Tokyo 2025 World Athletics Championships (13- 21 September 2025) to the World Athletics Road Running Championships in Copenhagen (19-20 September 2026), resulting in an overall data collection period of one year. No other changes have been made to the study objectives, eligibility criteria, survey content, recruitment procedures, outcome measures, or statistical analysis plan..

## REFERENCES

1 Elliott-Sale KJ, Ackerman KE, Lebrun CM, et al. Feminae: an international multisite innovative project for female athletes. BMJ Open Sport Exerc Med. 2023;9:e001675. doi: 10.1136/bmjsem-2023-001675

2 Giagio S, Salvioli S, Pillastrini P, et al. Sport and pelvic floor dysfunction in male and female athletes: A scoping review. Neurourol Urodyn. 2020;40:55–64. doi: 10.1002/nau.24564

3 Giagio S, Innocenti T, Pillastrini P, et al. What is known from the existing literature about the available interventions for pelvic floor dysfunction among female athletes? A scoping review. Neurourol Urodyn. 2022;41:573–84. doi: 10.1002/nau.24883

4 Donnelly GM, Bø K, Forner LB, et al. Up for the tackle? The pelvic floor and rugby. A review. Eur J Sport Sci. 2024;24:1719–34. doi: 10.1002/ejsc.12121

5 Moore I, Perkins J, Donnelly G. World Rugby Return to Rugby Postpartum Guidelines. Published Online First: February 2024. doi: 10.25401/cardiffmet.24759906.v2

6 Moore IS, Crossley KM, Bo K, et al. Female athlete health domains: a supplement to the International Olympic Committee consensus statement on methods for recording and reporting epidemiological data on injury and illness in sport. Br J Sports Med. 2023;57:bjsports-2022-106620. doi: 10.1136/bjsports-2022-106620

7 Rodríguez-López ES, Calvo-Moreno SO,Basas-García Á, et al. Prevalence of urinary incontinence among elite athletes of both sexes. J Sci Med Sport. 2021;24:338–44. doi: 10.1016/j.jsams.2020.09.017

8 Almeida MBA, Barra AA, Saltiel F, et al. Urinary incontinence and other pelvic floor dysfunctions in female athletes in Brazil: A cross-sectional study. Scand J Med Sci Sports. 2016;26:1109–16. doi: 10.1111/sms.12546

9 Teixeira RV, Colla C, Sbruzzi G, et al. Prevalence of urinary incontinence in female athletes: a systematic review with meta-analysis. Int Urogynecology J. 2018;29:1717–25. doi: 10.1007/s00192-018-3651-1

10 Dakic JG, Hay-Smith J, Cook J, et al. Screening for pelvic floor symptoms in exercising women: a survey of 636 health and exercise professionals. J Sci Med Sport. 2023;26:80–6. doi: 10.1016/j.jsams.2023.01.008

11 Ljungqvist A, Jenoure P, Engebretsen L, et al. The International Olympic Committee (IOC) Consensus Statement on periodic health evaluation of elite athletes March 2009. Br J Sports Med. 2009;43:631. doi: 10.1136/bjsm.2009.064394

12 Silvia G, Frederic G, Stephane B, et al. Current periodic health evaluation for athletes exclude pelvic floor health: are we neglecting an essential domain? J Sci Med Sport. Published Online First: 2025. doi: 10.1016/j.jsams.2025.07.001

13 Schulz JM, Pohlod L, Myers S, et al. Are female athlete specific health considerations being assessed and addressed in preparticipation examinations? A scoping review and proposed framework. J Sport Heal Sci. 2024;14:100981. doi: 10.1016/j.jshs.2024.100981

14 Cardoso AMB, Lima CRO de P, Ferreira CWS. Prevalence of urinary incontinence in high-impact sports athletes and their association with knowledge, attitude and practice about this dysfunction. Eur J Sport Sci. 2018;18:1405–12. doi: 10.1080/17461391.2018.1496146

15 Neels H, Tjalma WAA, Wyndaele J-J, et al. Knowledge of the pelvic floor in menopausal women and in peripartum women. J Phys Ther Sci. 2016;28:3020–9. doi: 10.1589/jpts.28.3020

16 Prudencio CB, Nava GT de A, Souza BR de, et al. Knowledge of pelvic floor disorders in young women: a cross-sectional study. Fisioter em Mov. 2022;35:e35607. doi: 10.1590/fm.2022.35607

17 Pereira E de S, Ferreira AP de L, Almeida M de O, et al. Prevalence and factors associated with urinary incontinence in female crossfitters: A cross-sectional study. LUTS: Low Urin Tract Symptoms. 2022;14:281– 8. doi: 10.1111/luts.12437

18 Vico-Moreno E, Fernández-Domínguez JC, Romero-Franco N, et al. An online workshop to raise awareness of pelvic floor in track and field female athletes: a quasi-experimental study. Arch Gynecol Obstet. 2024;1–8. doi: 10.1007/s00404-024-07790-x

19 Bosch-Donate E, Vico-Moreno E, Fernández-Domínguez JC, et al. Symptomatology and knowledge regarding pelvic floor dysfunctions and influence of gender stereotypes in female athletes. Sci Rep.2024;14:11052. doi: 10.1038/s41598-024-61464-x

20 Kiliç BB, Akgül H, Timurtaş E, et al. Knowledge Level of Pelvic Floor and Pelvic Floor Disorders According to and Related Disorders According to Gender and Education Levels. Int J Disabil Sports Heal Sci. 2023;6:101–10. doi: 10.33438/ijdshs.1245528

21 Leeuw ED de, Hox JJ, Dillman DA. International Handbook of Survey Methodology. 1st ed. Routledge 2008.

